# Chinese and Italian COVID-19 outbreaks can be correctly described by a modified SIRD model

**DOI:** 10.1101/2020.03.19.20039388

**Authors:** Diego Caccavo

## Abstract

The COVID-19 disease is rapidly spreading in whole globe, affecting millions of people and pushing governments to take drastic measures to contain the outbreaks. The understanding of the dynamics of the epidemic is of great interest for the governments and health authorities that are facing COVID-19 outbreaks. The scarce presence of epidemiologic data, due to the still ongoing outbreaks, makes prediction difficult and mainly based on heuristic (fitting) models. However, these models with non-physical based parameters, can only give limited insight in the evolution of the outbreaks. In this work a SIRD compartmental model was developed to describe and predict the evolution of the Chinese and Italian outbreaks. Exploiting the similarities of the measures taken by the governments to contain the virus and of the total population number of Hubei province and Italy, the model was tuned on the Chinese outbreak (almost extinguished) and by perturbation the Italian outbreak was describe and predicted.

## 1. Introduction

The COVID-19 disease, caused by severe acute respiratory syndrome coronavirus 2 (SARS-CoV-2), has spread rapidly in the first months of the 2020 and the World Health Organization (WHO) on the 11^th^ of March declared a global pandemic [1]. COVID-19 symptoms are often (80%) mild [2], may appear 2-14 days after exposure (mean incubation period 5-6 days) [2], and include fever, cough and shortness of breath [3]. However the 13.8% of infected have severe disease and the 6.1% are critical (respiratory failure, septic shock, and/or multiple organ dysfunction/failure) [2]. Mild cases typically recover within two weeks, while those with severe or critical disease may take three to six weeks to recover. Among those who have died, the time from symptom onset to death has ranged from two to eight weeks [2]. In the severe and critical disease, the patient admission to intensive care unit (ICU) is necessary and the mechanical ventilation (artificial assistance to support breathing), may be required [4]. This special treatment is the bottle neck of most of the health care systems, which are not prepared to deal with the numbers of patients that the COVID-19 disease could produce. Therefore, measures of containment of the SARS-CoV-2 outbreaks are essential to not overstress the health care systems and to manage the pandemic.

The SARS-CoV-2 was firstly reported at the end of 2019 in the Wuhan city (Hubei province), China, reaching epidemic proportion at the end of January 2020. On the 23^rd^ of January the Chinese government imposed the lockdown in Wuhan and other cities in Hubei province in an effort to quarantine the center of a COVID-19 outbreak. These drastic measures, with almost 60 million of people under lockdown, were defined “unprecedented in public health history” by the WHO. Unfortunately, these measures were not able to stop the spreading of SARS-CoV-2, which caused several outbreaks around the globe.

Italy was one of the countries most affected by the SARS-CoV-2, reporting the first positive patient on the 20^th^ of February 2020, followed by an extensive (and unexpected) epidemic in the north of Italy, especially in the Lombardy region. The Italian government, similarly to the Chinese government, progressively from the 8^th^ of March to the 11^th^ of March 2020 imposed the lockdown in the whole nation.

These drastic measures, also adopted by other EU countries, apart from the health sector, have a dramatic impact on the global economy as well as on the lifestyle habits of millions of people. Epidemiologic descriptions and predictions could help in the understanding of the dynamic of the COVID-19 disease, supporting governments and health authorities in the resources allocation [5]. Most of the models proposed until now to describe the COVID-19 outbreaks, due to the scarcely data available of a still ongoing epidemic, are heuristic models. These models are “fitting” equation, exponential [5] or logistic [6], used to describe the infection data, whose parameters have scarce (or none) physical meaning and their use for prediction, especially at long term, should be avoided. On the contrary compartmental models could constitute the right trade-off between the need of having physical based parameters, the easiness of solution and the scarce presence of data in the evolving epidemic. Typically, the population of interest is subdivided into a small number of compartments based on infection status (e.g. susceptible, infectious or recovered) and the flows between these compartments are described by ordinary differential equations [7]. Aim of this work is to develop a compartmental model (SIRD) to describe first the Chinese COVID-19 epidemic and then, by perturbation of the model parameters, to describe and predict the Italian COVID-19 epidemic.

## 2. SIRD Model

In compartmental models the population under study is divided into compartments and, with assumption that every individual in the same compartment has the same characteristics, the movements of people within the compartments can be described. In particular with “SIRD” model a four compartments model is indicated, where “S” indicates the “Susceptible” compartment, “I” specifies the “Infected” compartment, “R” denotes the “Recovered” compartment and “D” identifies the “Death” compartment, as schematized in Figure 1.

**Figure 1.**
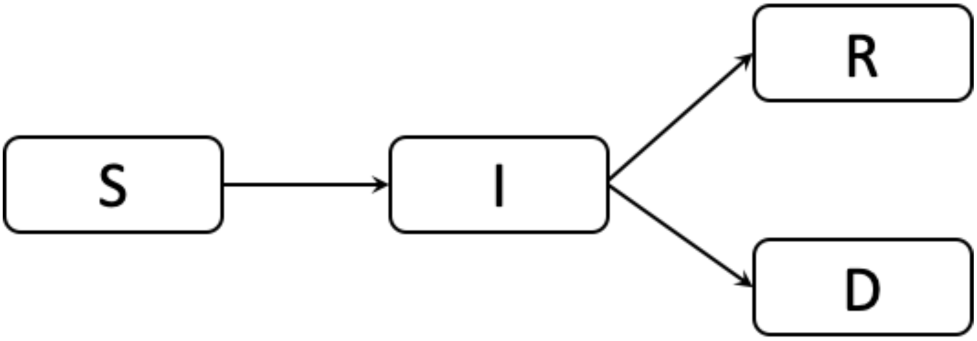
Schematization of the SIRD model. “S” indicates the “Susceptible” compartment, “I” specifies the “Infected” compartment, “R” denotes the “Recovered” compartment and “D” identifies the “Death” compartment.

It is assumed that the total population of reference is constant (N= 60 million), meaning that in the considered time period the rate of birth and death (including the one related to the COVID-19) are equal [8]. This is particularly true for low lethality diseases with very large outbreaks (like a whole nation). It is also assumed that recovered people cannot be re-infected [9]. The ordinary differential equations, with their initial conditions, describing the movements of people from one compartment to the other are reported in the eq. (1-4).

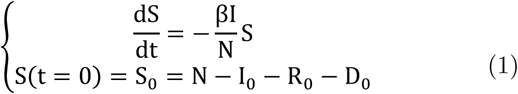

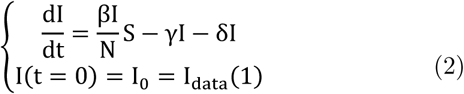

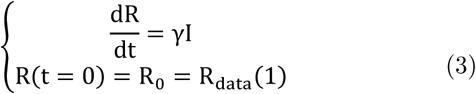

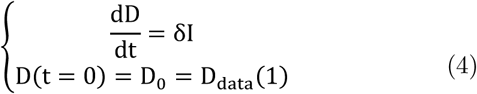

In eq. 1, *S* represents the number of people susceptible to the disease (total minus the already interested ones), *β* is transmission rate [1/day]. The probability to get infected is proportional to the product of the infected and the susceptible individuals, or better to the fraction of infected individuals (*I*/*N*) and the susceptible.

In eq. 2, *I* represents the number of infected people, whose initial value is taken from the literature data. Susceptible people get infected by the contact with other infected people (first term RHS), whereas infected people can recover (*γI*) with the kinetic *γ*, which is inversely proportional to the duration of the infection and directly proportional to the probability of recovery. Moreover, critical patients can die (*δI*) with the kinetic *δ*, which is inversely proportional to the average time between the infection and the death and directly proportional to the lethality of the disease. Eq. 3 and eq. 4 describe the time evolution of the number of recovered patients *R* and the death patients *D*. Actually, the four equations are not independent and the fourth one can be derived from the total balance on the population:

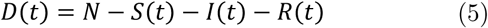

Despite very simple SIR (o SIRD) models are based on constant kinetics (*β, γ, δ*) to describe the epidemic evolution until the depletion of the susceptible people, the measures taken by the governments have had huge impact on the epidemic and therefore on its evolution kinetics.

### 2.1 β the kinetic of infection

The transmission rate *β* is most of the time used as fitting parameter to describe the epidemiologic data. In this case, with the lockdown protocol taken by the governments, the number of contacts per person per unit time can dramatically decrease during the epidemic. In this work the kinetic of infection was described with an exponential function:

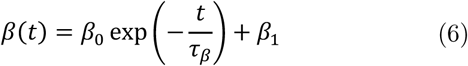

*β*_0_ (actually *β*_0_ + *β*_1_) is the initial (high) infection kinetics that decreases exponentially thanks to the lockdown measures. *β*_1_ is the infection kinetic at infinite time, which can be set to be zero due to a long enough period of people isolation. *τ*_*β*_ is the characteristic time of decrease, at *t =* 3 *τ*_*β*_ the kinetic of infection would be decreased of the 90%.

### 2.2 γ the kinetic of recovery

The kinetic of recovery *γ*, in case of a new disease like the COVID-19, may not be a constant. The health care system, the medical doctors with all their staff have to learn new therapeutic procedures, get used to treat patients with new symptoms etc. This means that initially the time of recovery will be longer, therefore the kinetic of recovery slower, until a new constant value is reached that should be inversely proportional to the average duration of the infection. In this work *γ* was modeled with a logistic function:

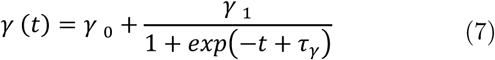

Where *γ*_0_ is the recovery kinetic at time zero, *γ*_0_+ *γ*_1_ is the recovery kinetic at regime, which is reached after *τ*_*γ*_ days of adaptation.

### 2.3 δ the kinetic of death

The kinetic of death *δ*, especially for new diseases, it is not constant and decreases with time, luckily. This is due to the adaptation of the pathogen, whose less lethal strains tend to be the most spread, as well as due to the development of new clinical treatments. Moreover, at the beginning of an epidemic usually the severe cases are observed first and the overall rate of dying appears to decrease when less severe cases are added. In this work an exponential decay function was used to describe this evolution:

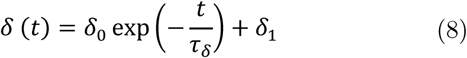

Where (*δ*_0_+*δ*_1_) is the initial death rate, which decreases substantially (90%) at *t =* 3 *τ*_δ_ and reaches a long-term lethality rate *δ*_1_.

### 2.4 ℛ: the reproduction number

In epidemiology, the basic reproduction number (ℛ_0_) of an infection can be thought of as the expected number of cases directly generated by one case in a population where all individuals are susceptible to infection [10]. When ℛ_0_> 1 the infection will be able to start spreading in a population, but not if ℛ_0_ < 1. Generally, the larger the value of ℛ_0_, the harder it is to control the epidemic. The reproduction number can be obtained from the SIR model dividing Eq 2 by the total population N [11]:

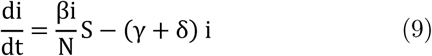

Where *i* is the normalized number of infected people (*I*/*N*). An epidemic occurs if the number of infected individuals increases, di/dt > 0, which means that the reproduction number (ℛ) has to be greater than the unity:

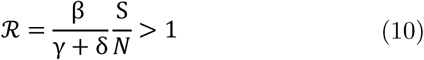

At the outset of an epidemic, nearly everyone is susceptible, so that S(0)/N∼1, but S(*t*)/N∼1 is also true for very large population with relatively low percentage of total cases:

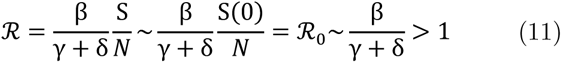

If the kinetic of infection β is higher than the sum of the kinetic of recovery *γ* and death δ, than the outbreak is epidemic, with ℛ_0_>1. On the contrary if β is small compared to the *γ* + δ than ℛ_0_<1 and the outbreak is destined to extinguish.

### 2.5 Implementation and optimization

The model was implemented and solved in MATLAB 2019b, with the ode45 solver, based on an explicit Runge-Kutta (4,5) formula. The optimization of the model parameters to describe the experimental results was performed via the minimization of the residuals (objective function) between the modeling and epidemiologic data. In particular, since the least squares approaches relies on the fitting of independent data, the use of the cumulative data should be avoided: cumulative incidence data are highly correlated. On the contrary the use of the daily new infection, daily recovery and daily deaths (incident cases) should be preferred, since these data are independent. However, these data are more scattered with respect to cumulative data and the ordinary least squares (OLS) which is highly sensitive to outliers, could generate misleading results. To overcome this potential pitfall, a robust regression method, based on the bisquare M-estimator was implemented.

Briefly, being *x*_*i*_ and 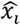 the modeling and the experimental data at a certain time point *i*, and being 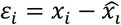 the residual, the bisquare robust fitting algorithm recalculates the residual at the time point according to eq. 9:

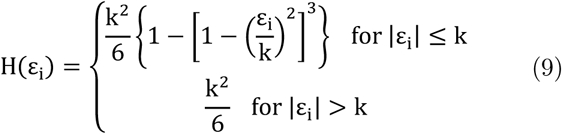

Where ***k =* 4**.**685 × MAD**(**ε**), where **MAD** is the mean absolute deviation of the values in the residuals [12]. With this approach the effects of outliers (|**ε**_+_| > **k**) is smoothed out. The sum of the **H**(**ε**_+_) constitutes the objective function to describe the goodness of fit between the model and the epidemiologic data. Since three independent data sets were used (daily new infection, daily recovery and daily deaths), the problem translates in a multi-objective function minimization. In this works the global objective function, weighted sum of the single objective functions, were minimized. The weights, needed to compensate the different order of magnitude between the single objective functions, were empirically chosen to be 1, 10 and 100 for the daily new infection, daily recovery and daily deaths residuals, respectively.

The global objective function was minimized with application of gradient-based method (fmincon) first, followed by a step of minimization with a gradient-free algorithm (pattern-search), followed by a third step of gradient-based method. The use of these three steps was necessary to reach global minima. The model describing the Italian outbreak is available at the reference [13].

## 3. Epidemiologic data

The Hubei outbreak can be considered a closed system since the lockdown measure, which isolated the province from the rest of the nation, coincide with starting point of availability of data. On the contrary the Italian outbreak could be approached in several ways, from the regional to the national level. However, considering at the beginning of the outbreak the high internal mobility (of people and wares) and the delayed lockdown, in this work the whole nation was considered a closed system.

The epidemiologic data were downloaded (automatically by the developed MATLAB script) from the open source repository operated by the Johns Hopkins University Center for Systems Science and Engineering (JHU CSSE) [14] for the Chinese (Hubei) outbreak and from the official open source repository operated by the Italian Department of Civil Protection on the official data provided by the Italian Ministry of Health [15]. However, the data reported in [14] for the Italian case are retrieved from [15], making the use of both the databases interchangeable.

The data regard the total number of cases, the recovered cases and the death cases. The number of infected can be estimated by the difference of the total number of cases minus the recovered and the death cases; all these data constitute the cumulative cases, (Figure 2A) and left side of Figure 2 B-D)). In particular for Figure 2B) the “cumulative” correspond to the total number of people that belong to the infected compartment (Figure 1): people that have been infected but are not recovered or death yet.

**Figure 2.**
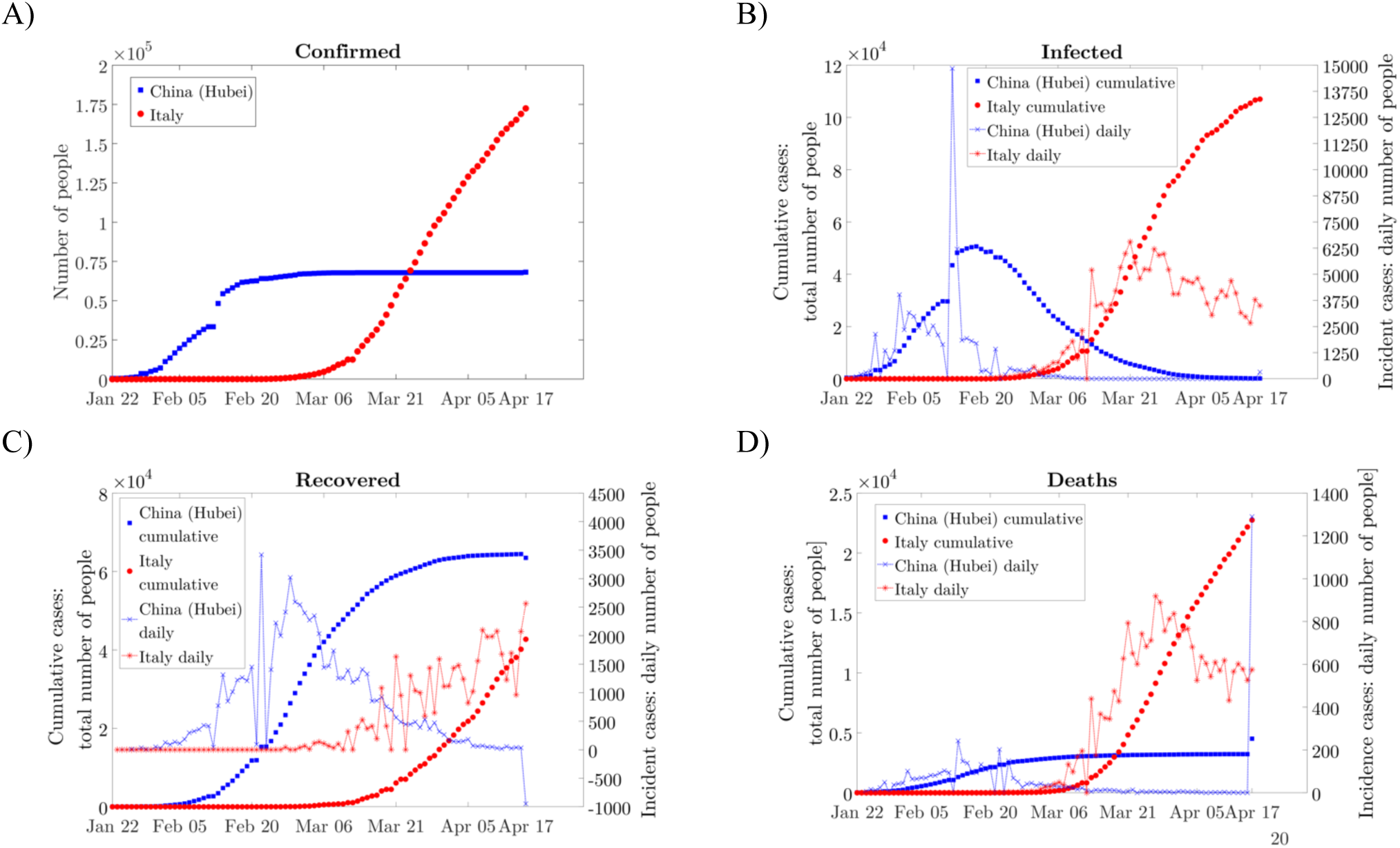
A) Confirmed cases. B) Infected people, “cumulative” cases on the left, incident cases on the right side. In this case the “cumulative” cases represent the total number of people infected but not recovered or death, yet. C) The number of recovered patients, cumulative cases on the left, incident cases on the right side. D) The number of dead patients, cumulative cases on the left, incident cases on the right side.

It is also useful to obtain the incident cases (new daily cases) which for the recovered people and the deaths is the derivative of their respective cumulative cases, whereas to obtain the incident infection the derivative of the total cases Figure 2 A) has to be performed.

From the experimental data this can be done calculating the incremental ratio of the cumulative cases. Being the minimum time span equal to 1 day, in this case the incremental ratio is equal to the difference between two adjacent points of the cumulative curves. The incident cases are reported on the right side of Figure 2 B-D).

As it can be seen the Chinese (Hubei) outbreak has been suppressed, with the peak of “cumulative” infected people around the 18^th^ of February. The spike recorded on the 13^th^ of February in the number of new infections was explained considering the inclusion in the counting of the cases individuated by clinical diagnosis (trough CT scan in the previous days) along with the cases individuated by lab tests. According to the released data at the end of the outbreak less than 70,000 people were infected by the SARS-CoV-2, and the 95% recovered. However, the Chinese government is recalculating the cases, as it can be seen from the last points in Figure 2 C-D): the deaths were increased of 1,290 people and the recovered decreased of 910 units (on the 17^th^ of April), reducing the recovery percentage at the 93%.

The Italian outbreak after the initial fast-growing phase, has reached the peak of incident infection at the end of March but with numbers way higher than the Hubei outbreak. On the 17^th^ of April 172,400 cases of COVID-19 were reported with 22,740 deaths and “only” 42,730 recovered patients. There is a strong debate on the problem of underestimating cases, especially related to the Chinese data but, also related to the impossibility of tracking worldwide (also in Italy) the SARS-CoV-2 infections that produce COVID-19 with mild symptoms. This could generate biased epidemiologic data (“Iceberg Phenomenon”) dominated by the most severe cases. Keeping in mind this problem, whose solution goes beyond the aim of the present work, the modeling results obtained from the use of these (biased) epidemiologic data will be critically discusses.

## 4. Results and discussion

The Chinese outbreak of COVID-19, at the end of its development, can be used to understand the dynamic of COVID-19 disease. The model parameters obtained for the description of the Chinese outbreak were perturbated to describe the initial Italian outbreak and to predict its evolution.

### 4.1 Chinese COVID-19 epidemic

The SIRD model results for the Chinese COVID-19 outbreak are reported in Figure 3. The model parameters (8 parameters) were fitted on the experimental data, obtaining a satisfactorily description of the epidemiologic data published so far. It is interesting to analyzed the value of the obtained modeling parameters, reported in Table 1 and plotted in Figure 4 A) to make some physical considerations. The transmission rate decreased with a characteristic time *τ*_*β*_=10.25 [day]. It is worth to note that the epidemiologic data are referred to the lockdown day -1. Since than the average number of contacts per person decreased exponentially, due to the lockdown, and diminished of 90% after almost 30 days (3*τ*_*β*_).

**Table 1.** Model parameters obtained from a best fit optimization.

| Parameter | China | Italy |
| --- | --- | --- |
| $\beta_0$ | 0.553 [1/day] | 0.256 [1/day] |
| $\tau_\beta$ | 10.25 [day] | 14.39 [day] |
| $\gamma_0$ | 0.015 [1/day] | 0.017 [1/day] |
| $\gamma_1$ | 0.061 [1/day] | - |
| $\tau_\gamma$ | 32.41 [day] | - |
| $\delta_0$ | 0.009 [1/day] | 0.024 [1/day] |
| $\delta_1$ | $6.6 \times 10^{-4}$ [1/day] | - |
| $\tau_\delta$ | 14.892 [day] | 21.6 [day] |

**Figure 3.**
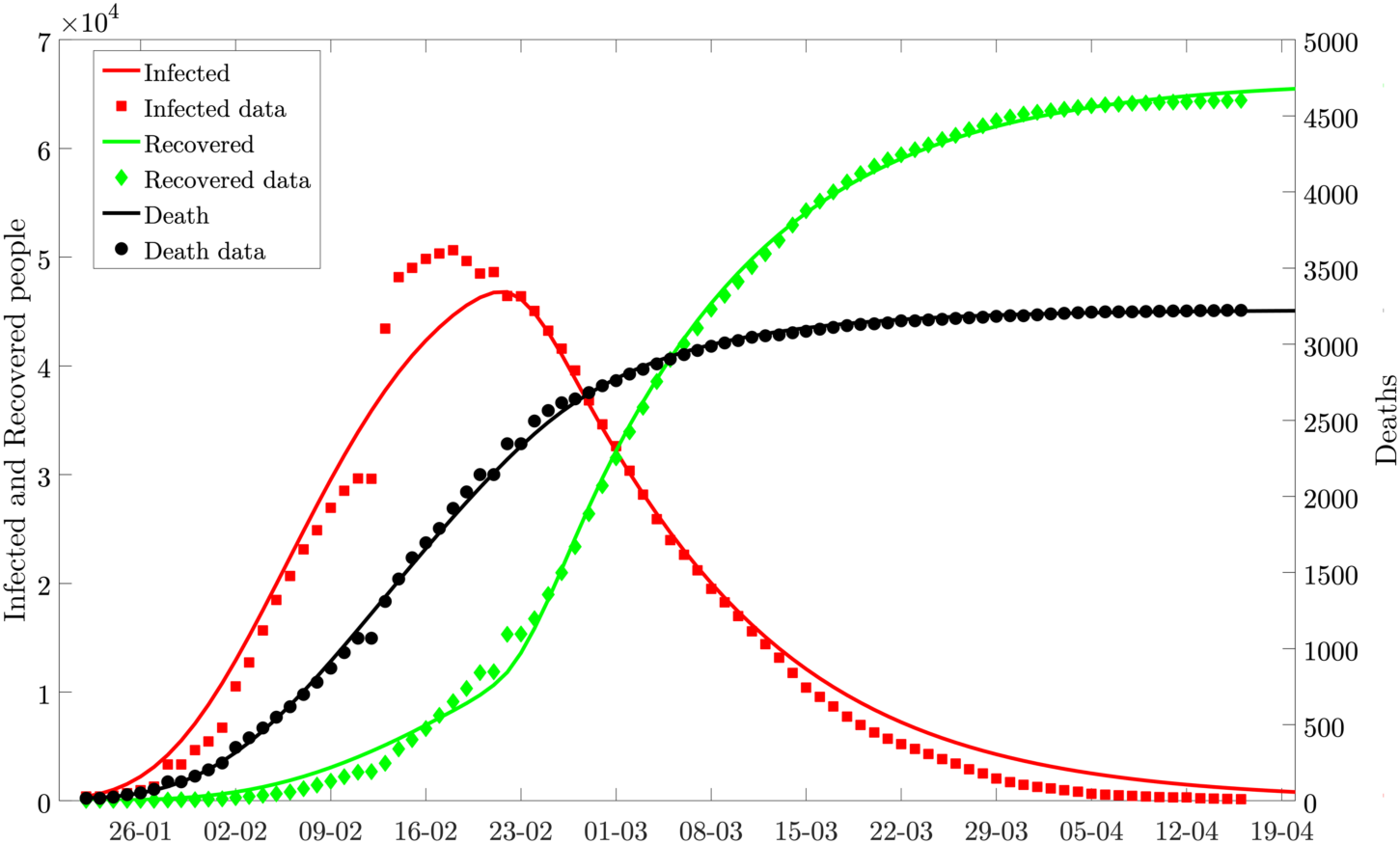
Chinese COVID-19 epidemiologic data descripted and predicted (at time longer than the data points) by the results of the SIRD model

**Figure 4.**
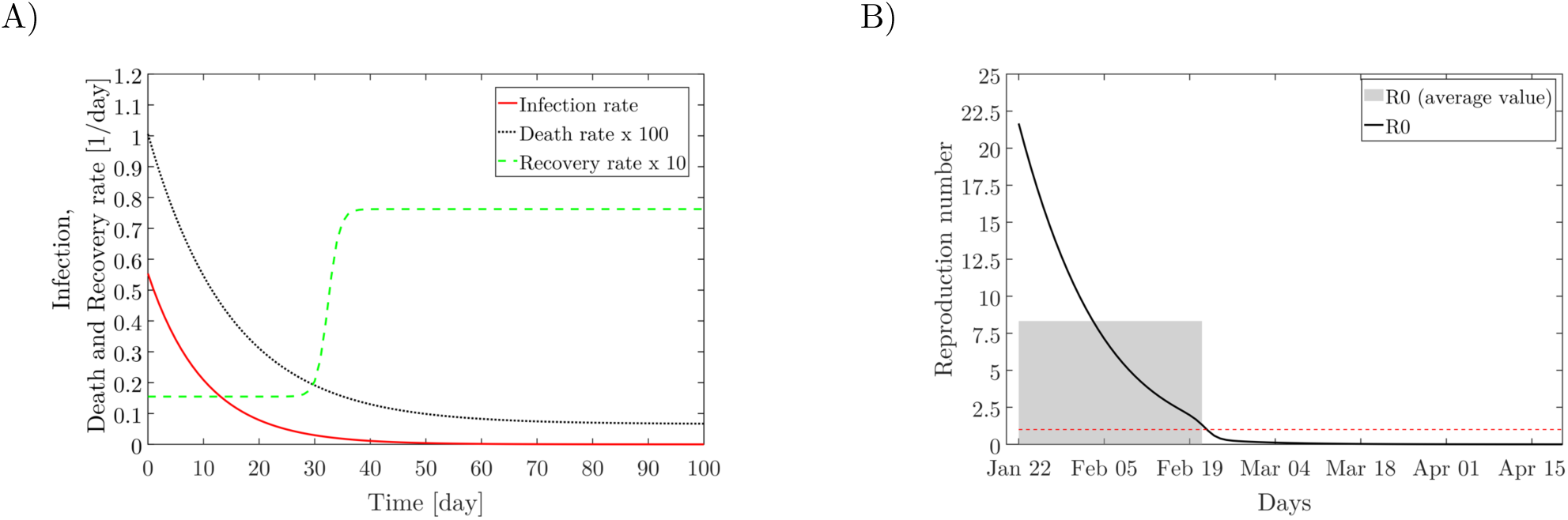
A) Infection, recovery and death kinetics for the Chinese (Hubei) outbreak obtained from the SIRD model. The death rate and the recovery rate have been multiplied by 100 and 10 respectively. B) The basic reproduction number evolution (black solid line), with the grey filled area the average reproduction number value (from the beginning of the epidemic until the decrease of ℛ_0_ at 1) is reported: <ℛ_0_>=8.3.

This demonstrate, along with the experimental date itself, that it required 30 days after the lockdown to substantially slow down the spreading of the virus.

The parameters referred to the kinetic of recovery *γ* _0_, *γ* _1_, *τ*_*γ*_ can give information on time of recovery (*d*_*recovery*_). Actually 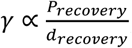, therefore considering the probability of recovery (*P*_*recovery*_) of 90% (R/(R+I+D)), the recovery time at regime (after 30 days (*τ*_*γ*_)) was *d*_*recovery*_*=* 12 days.

The parameters related to the kinetic of death, analogously can be used to obtain information about the average time between the symptoms and the death. Considering the death probability of 4.5% (D/(R+I+D)), the average death time was progressively increased from 5 days in the first week (22-28^th^ January) to 9 days considering the first 3 week, until reaching two months (meaning a lethality close to zero) at the end of the epidemic. This result can be explained considering that at the beginning of an epidemic the most severe cases are reported, displaying higher lethality, which tends to decrease when new cases less severe are added. Moreover it can be explained considering that the treatments were improved as well as considering a parameter not directly accounted in this model: the relation between lethality and patience age [16]. Indeed the lockdown measures may be particular effective for old people (>60 years) and for people with more comorbidities, who form the group of patients in which the COVID-19 lethality is the highest.

In Figure 4 B) the reproduction number for the Chinese outbreak is reported. With the black solid line the reproduction number calculated according to Eq. 11 is reported. The initial high value is mainly driven by the infection kinetic, which was particularly high in the province of Hubei. However, the lockdown measures were quite effective reducing the reproduction number to the unity in a month. The average value <ℛ_0_> during the epidemic (integral mean), reported with the gray area in Figure 4 B) was 8.3, meaning that in average a single sick person infected 8.3 people. This value is higher than the average of 3.28 reported in [17], where ℛ_0_ derived from different methods (in different works) were reported for the Chinese outbreaks. Actually, looking at the Huberi province [18] reported and ℛ_0_ of 6.5, [19] in some scenario reported and ℛ_0_ of 7.09 (90% CI: 5.84-8.35). In the initial stage of the epidemic on board of the Diamond Princess cruise ship the reproduction number was calculated to be 14.8 [20], lowered later thanks to the isolation and the quarantine protocol on board.

### 4.2 Italian COVID-19 epidemic

The Italian modeling of the outbreak was performed by perturbation of the Chinese parameters (starting points of the optimization). The modeling starts from the 26/02/2020, day in which the total cases in Italy were close to the cases in Hubei on the 22/01/2020 (around 400). The population of the Hubei province and Italy is also roughly the same: 60 million.

The main Italian lockdown was performed on the 11^th^ of March, when the cases were already 10590 (against the 438 on the 23^rd^ of January in China). To account for this delay, the equations for the description of the Italian outbreak were slightly adjusted for *β*(*t*) and *δ*(*t*).

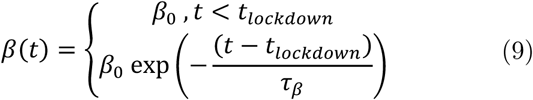

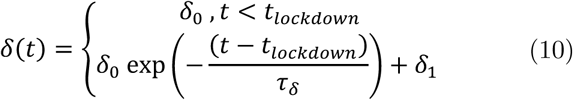

Considering a constant rate for the number of contacts per person and also a constant kinetic of death before the date of lockdown (between the 26^th^ of February and the 11^th^ of March). After the lockdown both the function decrease exponentially due to the isolation of people (reduction of *β*) and especially for categories that are at greatest risk of death (reduction of *δ*).

The modeling results obtained are reported in Figure 5.The epidemiologic data are well described with the optimization of only five adjustable parameters. Indeed, on the contrary of the Hubei outbreak, the model was able to describe the Italian epidemiologic data with only a parameter (*γ* _0_) for the kinetic of recovery. Moreover, on the experience of the Chinese outbreak, the long-term death kinetic was set to zero (δ_1_ *=* 0). The model parameters obtained are reported in Table 1 and in Figure 6 A). Most of the kinetic data are in agreement with the findings of [21].

**Figure 5.**
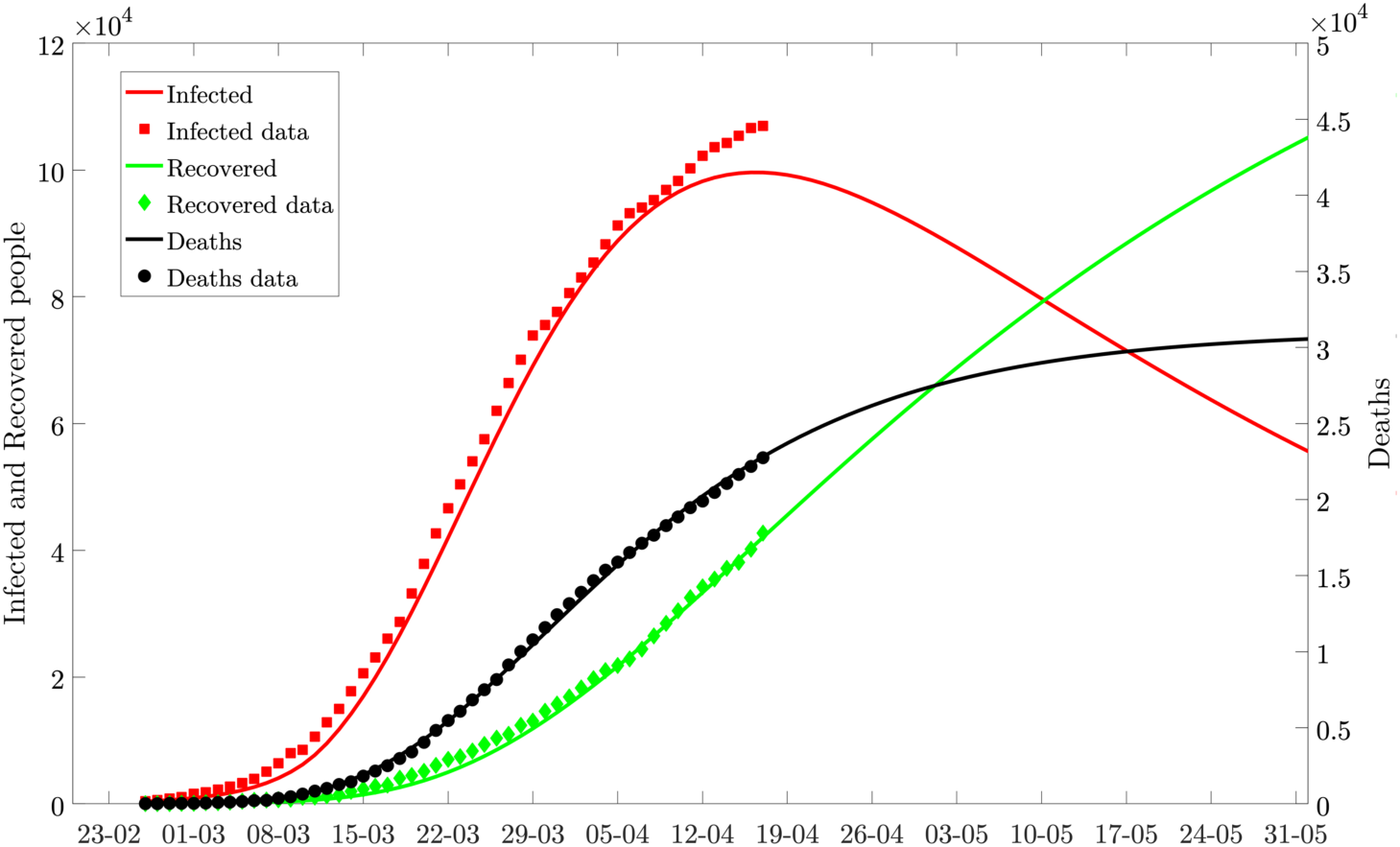
Italian COVID-19 epidemiologic data descripted and predicted (at time longer than the data points) by the results of the SIRD model

**Figure 6.**
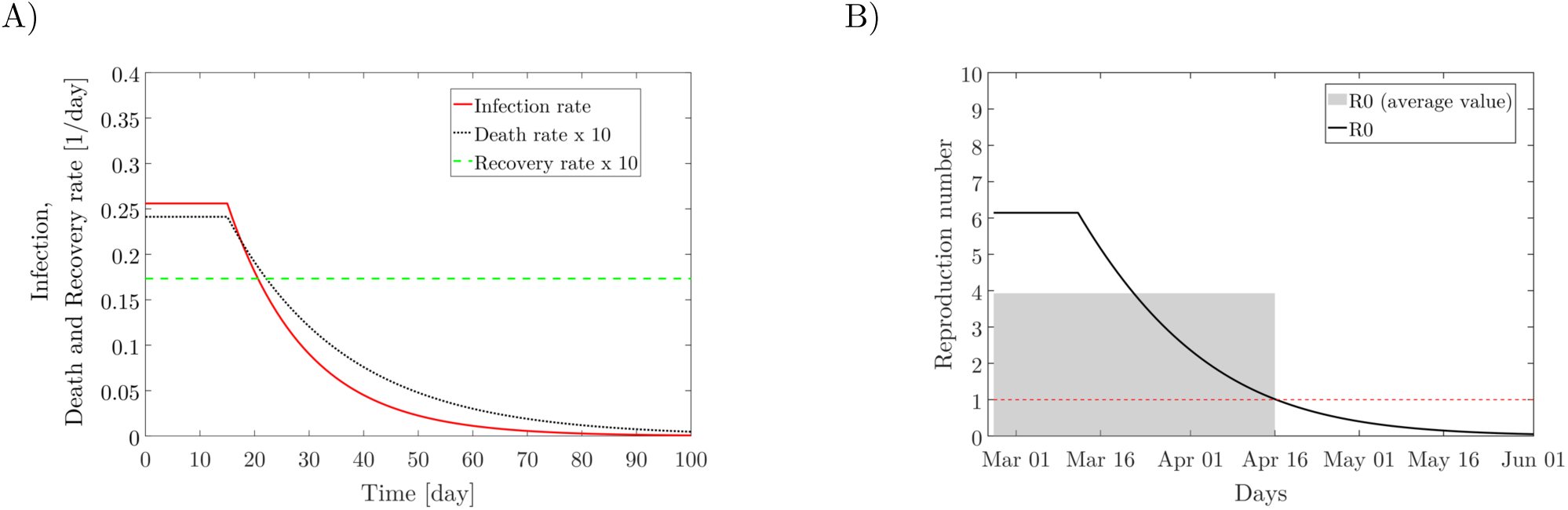
A) Infection, recovery and death kinetics for the Italian outbreak obtained from the SIRD model. B) The basic reproduction number evolution (black solid line), with the grey filled area the average reproduction number value (from the beginning of the epidemic until the decrease of ℛ_0_ at 1) is reported: <ℛ_0_>=3.9.

According to the model prediction the Italian outbreak on the 17^th^ of April is on the peak region, 42 days (3τ_β_) after the lockdown. It is interesting to note that despite the lower kinetic of infection (almost half of the Chinese), the retard in the application of the lockdown measures, which increased the number of circulating infected people before the lockdown, as well as the initial weaker restriction measures with respect to the Chinese ones, produced a long lasting epidemic.

As it can be seen in Figure 6 A) the kinetic of infection (red solid line), takes longer to decrease with respect to the Chinese outbreak Figure 4 A) and, despite starting from a value almost half of the Chinese, produced many more COVID-19 cases.

Moreover the Italian kinetic of recovery (green dashed line in Figure 6 A)), on the contrary of the Chinese, did not show (yet?) an increase, being able to describe the data with a constant (and very low) kinetic. This kinetic value generates a time of recovery *d*_*recovery*_ > 50 days. This result is in contrast with the average time of recovery from mild infection (2 weeks) [2], whereas can be representative of very severe infection (three to six weeks to recover) [2]. This seems to prove that the observed and reported cases in Italy, especially in the first period, were mostly severe cases. However, the addition of mild cases could produce in the following days and increase in the rate of recovery, generating also a quicker reduction of the infected patients.

The seriousness of the Italian COVID-19 cases can be seen also looking at the death kinetic, which is initially (*δ*_0_) 2.5 times greater than the Chinese and also slower in the decrease (higher *τ*_*δ*_), as it can be seen in Figure 6 A) (black dotted line). This caused many deaths in Italy (>20,000) and many other are expected, overcoming 30,000 cases. The higher lethality could be due to the higher median age of the Italian population (44.3 years) with respect to the Chinese population (37.4 years) [22]. However further studies should be performed to investigate such a high lethality, i.e. in [23] it is suggested that the reasons could be found in the Italian extensive intergenerational contacts, which are supported by a high degree of residential proximity between adult children and their parents. Other reasons, such as the careless management of the elderly in nursing homes is an ongoing political and criminal debate.

Such severe impact of the virus SARS-CoV-2 on the Italian would not have been expected looking at the initial reproduction number (Figure 6 B)). The lower infection kinetic recorded in Italy generated an initial ℛ_0_ of 6 (way lower than the Chinese). However, ℛ_0_ decreased slower due to the retarded lockdown (constant value in (Figure 6 B)) and due to the not increasing recovery rate. The average <ℛ_0_> in Italy was calculated to be 3.9, close to the ℛ_0_ range of [2.13 - 3.33] reported in [24].

Such difference in the “force of infection” could be explained considering that the mains outbreaks in Italy have been developed in small cities, whereas in the Hubei province the principal outbreak was in the metropolitan city of Wuhan (11 million of people).

The Italian outbreak, still in the peak region, has showed stable death and recovery kinetic over the past 30 days, indeed fitting backward in time the Italian data the parameters *γ* _0_, *δ*_0_, *τ*_*δ*_ result quite stable. The most important parameter, which causes the retardation of the decreasing phase of the epidemic is the characteristic time *τ*_*β*_ in the infection kinetic. The higher *τ*_*β*_, the longer it will take to reduce the number of cases (the longer it will take to reach ℛ_0_<1). In Figure 7 the impact of the variation of *τ*_*β*_ is reported on incident cases. With the blue solid thick line the “best fit” value (used in this work) is reported. However, as it can be seen from the experimental data, the epidemiologic data are prone to be describe with other characteristic time of infection *τ*_*β*_, especially higher than the present “best fit” value. Therefore, depending on the new infection of the next days the peak could slightly translate, however with limited impact on the deaths count.

**Figure 7.**
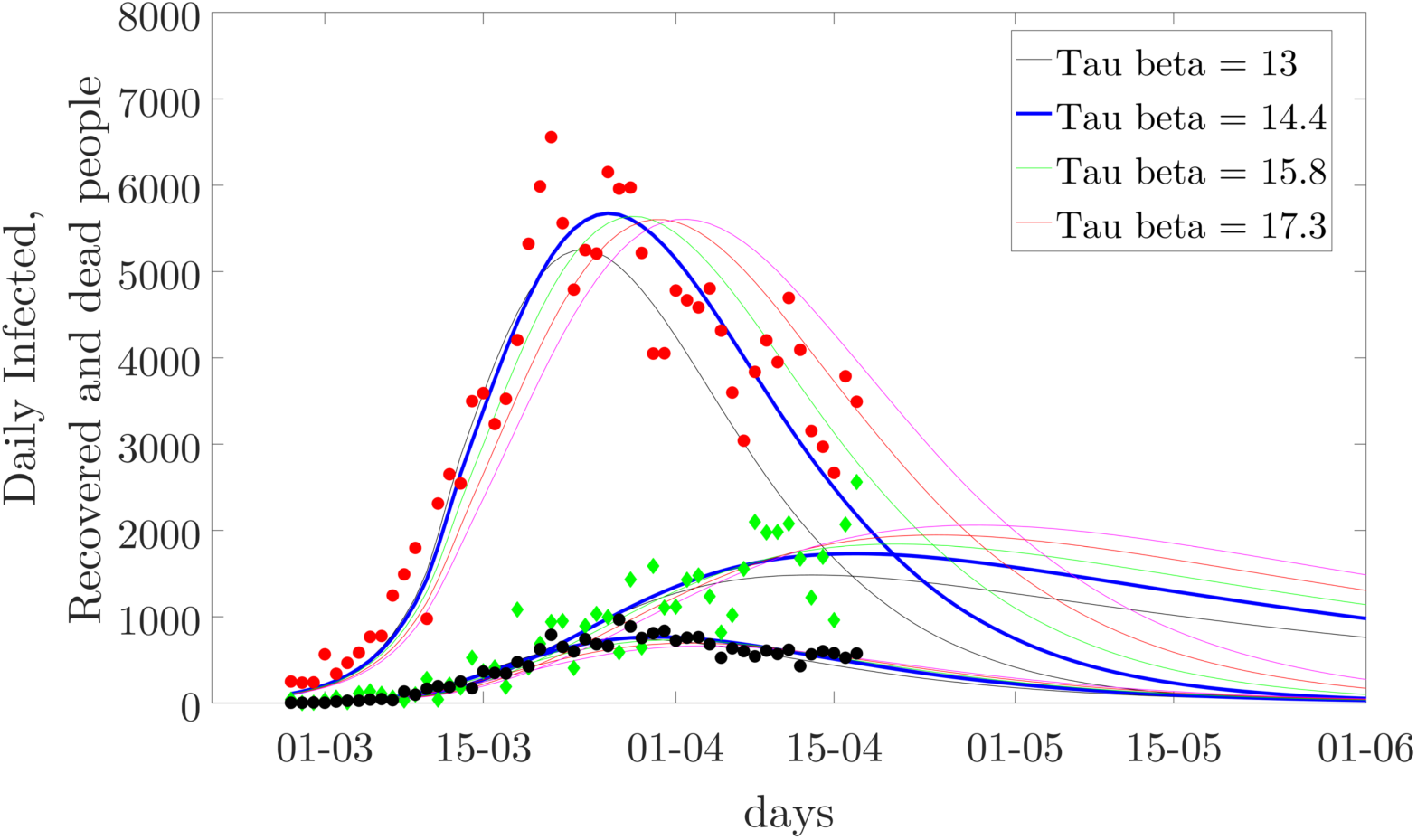
Parametric sweep of τ_*β*_. The incident infection (red circle), recovery (green diamonds) and death cases (black circle) are reported. With the solid lines the modeling results at different value of τ_*β*_ are reported.

### Conclusions

The understanding of the dynamic of the epidemic could be useful to take decisions in time, preparing hospitals to deal with the infection peak. The ongoing outbreaks are often described with simple fitting models, i.e. exponential functions (and/or sigma-shaped curves such as Logistic or Gompertz), whereas in this work, a SIRD compartmental model was developed and used to describe and predict the evolution of the Chinese (Hubei) and of the Italian outbreaks. The advantages of SIRD models with respect to heuristic models reside mainly in the physical-based SIRD parameters, which can give useful insight and/or can be properly chosen on the basis of physical-based assumptions.

The present SIRD model was able to describe the outbreaks, obtaining the kinetic of infection, the kinetic of recovery as well as the kinetic of death. The Italian best fit initial kinetic of infection was lower (half) than the Chinese one, probably because the Italian outbreaks are mainly based in small hospitals/cities, whereas the main Chinese outbreak was in the metropolitan city of Wuhan. However, the decrease time of this kinetic in the Italian outbreak was slower than in the Chinese outbreak, probably due to the delayed lockdown and initial weaker restrictions. Therefore, despite a lower average ℛ_0_, 3.9 in Italy versus 8.3 in Hubei, the longer Italian epidemic produced much more COVID-19 cases.

The Italian epidemic showed a higher death kinetic, causing at the end of infection 10 times the Chinese deaths (modeling results). This, along with a very slow recovery kinetic, proved that most of the reported cases in Italy are related to severe infections, therefore the epidemiologic data could be highly underestimated in the total number of cases as well as in the recovered cases.

On the contrary, according to the data released so far, the Draconian lockdown measures taken by the Chinese government have been more effective, extinguishing in 1 month an outbreak that was potentially more dangerous than the Italian one.

## Data Availability

The data that support the findings of this study are openly available from the 2019 Novel Coronavirus COVID-19 (2019-nCoV) Data Repository by Johns Hopkins CSSE

https://github.com/CSSEGISandData/COVID-19

https://it.mathworks.com/matlabcentral/fileexchange/74838-sird-model-for-covid-19-outbreaks

## Acknowledgements

Thanks to professor Gaetano Lamberti for the constructive discussions and for the pre-reviewing of the manuscript.

## References

1. WHO. WHO Director-General’s opening remarks at the media briefing on COVID-19 - 11 March 2020. 2020; Available from: https://www.who.int/dg/speeches/detail/who-director-general-s-opening-remarks-at-the-media-briefing-on-covid-1911-march-2020.

2. WHO. Report of the WHO-China Joint Mission on Coronavirus Disease 2019 (COVID-19). 2020; Available from: https://www.who.int/docs/default-source/coronaviruse/who-china-joint-mission-on-covid-19-final-report.pdf.

3. CDCP. Centers for Disease Control and Prevention. Coronavirus symptoms and diagnosis. 2020; Available from: https://www.cdc.gov/coronavirus/2019-ncov/symptoms-testing/symptoms.html.

4. Guan, W.-j., et al., Clinical Characteristics of Coronavirus Disease 2019 in China. New England Journal of Medicine, 2020.

5. Remuzzi, A. and G. Remuzzi, COVID-19 and Italy: what next? The Lancet, 2020

6. Batista, M., Estimation of the final size of the second phase of the coronavirus epidemic by the logistic model. medRxiv, 2020: p. 2020.03.11.20024901.

7. Lloyd, A.L. and S. Valeika, Network models in epidemiology: an overview, in Complex population dynamics: nonlinear modeling in ecology, epidemiology and genetics. 2007, World Scientific. p. 189–214.

8. Brauer, F., P.d. Driessche, and J. Wu, Lecture notes in mathematical epidemiology. 2008: Berlin, Germany: Springer.

9. Bao, L., et al., Reinfection could not occur in SARS-CoV-2 infected rhesus macaques. bioRxiv, 2020: p. 2020.03.13.990226.

10. Fraser, C., et al., Pandemic Potential of a Strain of Influenza A (H1N1): Early Findings. Science, 2009. 324(5934): p. 1557.

11. Jones, J.H., Notes on R0. 2007, Stanford University.

12. Andersen, R. and S. Publications, Modern Methods for Robust Regression. 2008: SAGE Publications.

13. Caccavo, D. SIRD model for COVID-19 outbreaks 2020; Available from: https://www.mathworks.com/matlabcentral/fileexchange/74838-sird-model-for-covid-19-outbreaks.

14. Johns Hopkins University Center for Systems Science and Engineering. 2019 Novel Coronavirus COVID-19 (2019-nCoV) Data Repository by Johns Hopkins CSSE. 2020; Available from: https://github.com/CSSEGISandData/COVID-19.

15. Italian Department of Civil Protection. Dati COVID-19 Italia. 2020; Available from: https://github.com/pcm-dpc/COVID-19.

16. Zhonghua, L.X., Bing; Xue; Za Zhi The Novel Coronavirus Pneumonia Emergency Response Epidemiology Team. The Epidemiological Characteristics of an Outbreak of 2019 Novel Coronavirus Diseases (COVID-19). China CDC Weekly, 2020, 2(8): 113-122, 2020.

17. Liu, Y., et al., The reproductive number of COVID-19 is higher compared to SARS coronavirus. Journal of Travel Medicine, 2020. 27(2).

18. Shen, M., et al., Modelling the epidemic trend of the 2019 novel coronavirus outbreak in China. bioRxiv, 2020: p. 2020.01.23.916726.

19. Anastassopoulou, C., et al., Data-based analysis, modelling and forecasting of the COVID-19 outbreak. JPLOS ONE, 2020. 15(3): p. e0230405.

20. Rocklöv, J., H. Sjödin, and A. Wilder-Smith, COVID-19 outbreak on the Diamond Princess cruise ship: estimating the epidemic potential and effectiveness of public health countermeasures. Journal of Travel Medicine, 2020.

21. Calafiore, G.C., C. Novara, and C. Possieri, A Modified SIR Model for the COVID-19 Contagion in Italy. arXiv preprint arXiv:2003.14391, 2020.

22. WHO. Population Data by country (Recent years) Available from: https://apps.who.int/gho/data/view.main.POP2040

23. Dowd, J.B., et al., Demographic science aids in understanding the spread and fatality rates of COVID-19. medRxiv, 2020: p. 2020.03.15.20036293.

24. Riccardo, F., et al., Epidemiological characteristics of COVID-19 cases in Italy and estimates of the reproductive numbers one month into the epidemic. medRxiv, 2020: p. 2020.04.08.20056861.

